# The optimal second arterial graft and sex differences in coronary bypass surgery: 10-year national registry results

**DOI:** 10.64898/2026.04.04.26350161

**Authors:** Sophie H.Q. Beukers, Edgar J. Daeter, Hans Kelder, Saskia Houterman, Geoffrey T.L. Kloppenburg, the Cardiothoracic Surgery Registration Committee of the Netherlands Heart Registration

## Abstract

**Background:** Disparities between sexes in mortality and morbidity after coronary artery bypass grafting remain incompletely understood. Multi-arterial grafting demonstrates superior outcome compared to single arterial grafting, although the optimal type of a second arterial graft and possible sex-dependent differences in grafting strategy have not been elucidated. We aim to determine whether the right internal thoracic artery or the radial artery is the optimal second arterial graft.

**Methods:** We analyzed data from 14,196 patients undergoing primary isolated coronary artery bypass grafting with the left internal thoracic artery and either right internal thoracic artery or radial artery between 2013 and 2022 from the Netherlands Heart Registration. Patients were stratified by sex and type of second arterial graft. Inverse probability treatment weighting was used to balance baseline characteristics. The primary outcome was long-term mortality. Secondary outcomes included short-term complications and repeat revascularization.

**Results:** In both sexes, the choice of second arterial graft did not significantly impact long-term survival. Postoperative arrhythmias were more prevalent in both sexes following right internal thoracic artery use (p<0.001). The radial artery was associated with higher rate of repeat revascularization in men (p=0.044 at 5 years follow-up) and more cerebrovascular accidents in women (0.9% vs 0.2%, p=0.028).

**Conclusion:** The choice of second arterial graft did not affect long-term survival in either sex. The radial artery was associated with an increased risk of repeat revascularization in men and more cerebrovascular accidents in women. These results underscore the need for further research in the field of sex-specific considerations in operative strategy.

## INTRODUCTION

The benefits of multi-arterial grafting (MAG) over single-arterial grafting in coronary artery bypass grafting (CABG) have been well established, including improved long-term survival and reduced incidence of major adverse cardiac events, irrespective of patient sex [1–6]. Left internal thoracic artery (LITA) to the left anterior descending coronary artery is considered the gold standard in CABG. The selection of a second graft depends on several factors, including comorbidity, stenosis severity of the target coronary artery, quality of graft vessels and surgeon expertise. Guidelines on myocardial revascularization recommend MAG with the LITA combined with either the radial artery (RA) or right internal thoracic artery (RITA) over saphenous vein grafts, unless contra-indicated [7,8].

The RAPCO-RITA trial – the only randomized controlled trial (RCT) to date comparing the RA to the RITA as the second arterial graft – revealed superior long-term patency and survival in the RA cohort [9]. Women are frequently underrepresented in clinical trials, complicating evidence-based decision-making for this population. Female sex has been independently associated with higher all-cause mortality and graft failure after CABG, resulting in increased rates of myocardial infarction and re-intervention [10]. Women undergoing CABG more often suffer from comorbidities such as diabetes mellitus, hypercholesterolemia, hypertension and peripheral arterial disease, which are risk factors for RA atherosclerosis and profound intimal hyperplasia [11–15]. The internal thoracic arteries are more resistant to deleterious effects of these comorbidities [13]. Consequently, women may face a higher risk of RA graft failure compared to men, potentially contributing to worse postoperative outcomes [10].

Evidence regarding the interaction between graft subtype and sex is limited. This raises the question whether women derive similar clinical benefits of the RA as the second arterial conduit compared to men. We compared the results of MAG in the Netherlands using RITA or the RA in both sexes, to help determine whether sex should be considered during preoperative planning of the grafting strategy in CABG.

## METHODS

### Data and ethics

Study protocol and data request were approved by the Netherlands Heart Registration (NHR). The NHR is a prospective registry that collects data on all procedures performed in cardiothoracic surgical centers across the Netherlands. The processes for data acquisition and quality assessment have been described previously [16,17]. A waiver for informed consent for analysis of the anonymized data of the NHR registry was obtained from the institutional review board Medical research Ethics Committees United (W19.270). The study was conducted in agreement with the principles of the Declaration of Helsinki. Local approval was obtained 12th of August 2025 (Z25.046).

### Population

Patients undergoing isolated multi-arterial CABG using the LITA and RITA or RA in the Netherlands between January 2013 and December 2022 were included. Exclusion criteria were prior cardiac surgery and patients receiving more than 2 arterial grafts. Patients were first categorized by sex and then stratified by the type of second arterial graft—RA or RITA.

### Outcome measures

Primary outcome was long-term all-cause mortality, with secondary outcomes long-term repeat revascularization and the incidence of short-term postoperative complications. Short-term complications during postoperative hospital stay included repeat cardiac surgery, cerebrovascular accidents (CVA), new-onset arrhythmias requiring intervention (cardioversion or medication; including atrial fibrillation) or death.

### Statistical analysis

Continuous values are presented as mean ± standard deviation, or median [interquartile range Q1-Q3], depending on distribution. Normal distribution was tested using the Quantile-Quantile plot and Kolmogorov-Smirnov test. Continuous variables were compared using the independent sample t-test when normally distributed, or using the Mann-Whitney U test when non-normally distributed. Categorical values were compared between subgroups using the Chi square test. An alpha level of 0.05 or less was considered statistically significant. All statistical analyses were performed using R version 4.4.2 (https://www.R-project.org/).

To compare RA with RITA treatment, we adjusted for measured confounding using inverse probability of treatment weighting (IPTW) based on propensity scores. The propensity scores were calculated using baseline variables: age, comorbidities, weight and height and procedural urgency. Comorbidities included were diabetes mellitus, estimated glomerular filtration rate, left ventricular ejection fraction (LVEF), peripheral arterial disease, chronic pulmonary disease, prior CVA and recent myocardial infarction (within 90 days prior to surgery). A standardized mean difference (SMD) <0.10 was considered a negligible imbalance between groups. Applying the weights was performed with the survey package for R (https://CRAN.R-project.org/package=survey).

Separate analyses were performed on females and males. Short-term postoperative complications were analyzed if <5% was missing, and the results table excludes missing values. Overall survival was assessed with the Kaplan-Meier method, comparing the curves between groups using the Log-rank test; we used Cox proportional hazard modelling to estimate the hazard ratios. To compare repeat revascularization between treatment cohorts, a competing risk analysis involving mortality and repeat revascularization was performed by means of a non-parametric cumulative incidence plot as implemented in the causalCmprsk package for R (https://CRAN.R-project.org/package=causalCmprsk).

## RESULTS

### Baseline characteristics

Between 2013 and 2022, 14470 patients underwent primary isolated CABG utilizing the LITA combined with either the RITA or the RA. Before adjustment, patients in the RA cohorts were younger and had a lower prevalence of most comorbidities than those in the RITA cohorts in both sexes, resulting in a lower Euroscore II (Table 1).

**Table 1.** Unadjusted baseline characteristics of the RA and RITA subgroups in men and women.

| Characteristics | Men | Women |
| --- | --- | --- |

|  | **RA**<br>N = 7,491 | **RITA**<br>N = 4,970 | **SMD** | **RA**<br>N = 1,117 | **RITA**<br>N = 892 | **SMD** |
| --- | --- | --- | --- | --- | --- | --- |
| Age (years) | 59 ± 8) | 61 ± 9 | 0.30 | 61 ± 10 | 65 ± 10 | 0.40 |
| Diabetes mellitus | 1,229 (17) | 1,206 (24) | 0.18 | 247 (23) | 311 (35) | 0.27 |
| Unknown | 212 | 2 | 0.24 | 37 | 0 | 0.26 |
| Height (cm) | 179 ± 7 | 179 ± 7 | 0.03 | 165 ± 8 | 165 ± 7 | 0.08 |
| Unknown | 366 | 192 | 0.05 | 64 | 41 | 0.05 |
| Weight (kg) | 87 ± 13 | 90 ± 15 | 0.24 | 73 ± 12 | 79 ± 15 | 0.40 |
| Unknown | 328 | 153 | 0.07 | 56 | 35 | 0.05 |
| Kidney function |  |  |  |  |  |  |
| eGFR ≥ 60 | 6,825 (91) | 4,305 (88) | 0.12 | 918 (83) | 674 (77) | 0.14 |
| eGFR 30-59 | 611 (8.2) | 562 (11) | 0.11 | 182 (16) | 191 (22) | 0.14 |
| eGFR 15-29 | 16 (0.2) | 30 (0.6) | 0.06 | 9 (0.8) | 12 (1.4) | 0.05 |
| eGFR < 15 | 8 (0.1) | 1 (<0.1) | 0.03 | 2 (0.2) | 0 (0) | 0.06 |
| Dialysis | 6 (<0.1) | 6 (0.1) | 0.01 | 1 (<0.1) | 0 (0) | 0.04 |
| Unknown | 25 | 66 | 0.11 | 5 | 15 | 0.12 |
| LVEF |  |  |  |  |  |  |
| [10%-30%) | 79 (1.1) | 117 (2.4) | 0.10 | 16 (1.5) | 19 (2.1) | 0.05 |
| [30%-40%) | 109 (1.5) | 77 (1.6) | 0.01 | 20 (1.8) | 12 (1.3) | 0.04 |
| [40%-50%) | 1,024 (14) | 1015 (20) | 0.17 | 118 (11) | 125 (14) | 0.10 |
| [50%-60%) | 5,161 (71) | 3074 (62) | 0.18 | 792 (73) | 607 (68) | 0.11 |
| [60%-82%] | 940 (13) | 673 (14) | 0.02 | 138 (13) | 127 (14) | 0.05 |
| Unknown | 178 | 14 | 0.18 | 33 | 2 | 0.22 |
| Chronic pulmonary disease | 428 (5.7) | 391 (7.9) | 0.09 | 91 (8.1) | 96 (11) | 0.09 |
| Peripheral arterial disease | 520 (6.9) | 463 (9.3) | 0.09 | 100 (9.0) | 107 (12) | 0.10 |
| Neurological dysfunction | 65 (0.9) | 66 (1.3) | 0.04 | 17 (1.6) | 16 (1.8) | 0.01 |
| Unknown | 341 | 19 | 0.27 | 63 | 3 | 0.32 |
| Critical preoperative state | 64 (0.9) | 110 (2.2) | 0.11 | 14 (1.3) | 21 (2.4) | 0.08 |
| Recent myocardial infarction | 2,297 (31) | 1,675 (34) | 0.06 | 320 (29) | 301 (34) | 0.11 |
| Unknown | 8 | 4 | 0.01 | 2 | 0 | 0.06 |
| Euroscore II | 1.34 ± 1.30 | 1.63 ± 1.86 | 0.18 | 2.28 ± 2.56 | 2.72 ± 2.78 | 0.16 |
| Unknown | 13 | 20 | 0.04 | 7 | 3 | 0.04 |
| Procedural urgency |  |  |  |  |  |  |
| Elective | 3,693 (50) | 2,464 (50) | 0.02 | 513 (47) | 448 (50) | 0.06 |
| Urgent | 3,520 (48) | 2,404 (48) | 0.01 | 537 (49) | 418 (47) | 0.05 |
| Emergent | 103 (1.4) | 91 (1.8) | 0.03 | 30 (2.8) | 24 (2.7) | 0.00 |
| Salvage | 6 (<0.1) | 10 (0.2) | 0.03 | 5 (0.5) | 2 (0.2) | 0.04 |
| Unknown | 169 | 1 | 0.21 | 32 | 0 | 0.24 |
| Prior CVA | 160 (2.7) | 165 (3.7) | 0.05 | 32 (3.7) | 36 (4.5) | 0.04 |
| Unknown | 1610 | 466 | 0.34 | 257 | 87 | 0.36 |

Mean ± standard deviation, number (percentage). Abbreviations: CVA, cerebrovascular
event; eGFR, estimated glomerular filtration rate; LVEF, left ventricular ejection fraction; RA,
radial artery; RITA, right internal thoracic artery; SMD, standardized mean difference.
difference; eGFR, estimated glomerular filtration rate; LVEF, left ventricular ejection fraction;
CVA, cerebrovascular event.

Following inverse probability treatment weighting adjustment, baseline characteristics were well-balanced between treatment cohorts. The majority of patients presented in an elective or urgent setting. The male population consisted of 11,694 patients, of which 7,204 (59.6%) received the RA and 4490 (40.4%) the RITA. Less than 10% had decreased kidney function, 20% suffered from diabetes mellitus, peripheral arterial disease was observed in approximately 8%, leading to a mean Euroscore II of 1.4%.

After adjustment, 1,783 female patients were included in the analysis, of which 1,009 (54.8%) received the RA, while 774 (45.2%) underwent the procedure with the RITA. In the female treatment cohorts, 4% had suffered a prior CVA, and kidney function was decreased in 20%. The mean Euroscore II was calculated to be 2.4% and did not differ statistically between treatment groups. A limited statistical difference was seen in weight between the RA and RITA cohorts (75±13kg vs 76±14kg, SMD 0.10) (Table 2).

**Table 2.** Baseline characteristics in men and women after inverse probability treatment weighting between the RA and RITA.

| Characteristics | Men |  |  | Women |  |  |
| --- | --- | --- | --- | --- | --- | --- |
|  | RA<br>N = 7,204 | RITA<br>N = 4,490 | SMD | RA<br>N = 1,009 | RITA<br>N = 774 | SMD |
| Age (years) | 59 ± 8 | 60 ± 9 | 0.05 | 62 ± 10 | 63 ± 10 | 0.09 |
| Diabetes mellitus | 1,343 (19) | 921 (21) | 0.04 | 254 (26) | 222 (29) | 0.07 |
| Unknown | 129 | 10 | 0.13 | 21 | 0 | 0.16 |
| Height (cm) | 179 ± 7 | 179 ± 7 | 0.01 | 165 ± 7 | 165 ± 7 | 0.03 |
| Unknown | 314 | 133 | 0.07 | 48 | 26 | 0.07 |
| Weight (kg) | 88 ± 13 | 89 ± 14 | 0.04 | 75 ± 13 | 76 ± 14 | 0.10 |
| Unknown | 273 | 109 | 0.07 | 42 | 22 | 0.07 |
| Kidney function |  |  |  |  |  |  |
| eGFR ≥ 60 | 6,475 (90) | 4,009 (90) | 0.01 | 810 (81) | 615 (80) | 0.01 |
| eGFR 30-59 | 657 (9.2) | 425 (9.5) | 0.01 | 182 (18) | 143 (19) | 0.01 |
| eGFR 15-29 | 14 (0.2) | 15 (0.3) | 0.02 | 9 (0.9) | 8 (1.0) | 0.01 |
| eGFR < 15 | 6 (<0.1) | 0 (<0.1) | 0.03 | 1 (0.1) | 0 (0) | 0.04 |
| Dialysis | 5 (<0.1) | 3 (<0.1) | 0.00 | 1 (0.1) | 0 (0) | 0.06 |
| Unknown | 47 | 37 | 0.02 | 5 | 9 | 0.06 |
| LVEF |  |  |  |  |  |  |
| [10%-30%) | 98 (1.4) | 78 (1.8) | 0.03 | 16 (1.6) | 15 (1.9) | 0.02 |
| [30%-40%) | 106 (1.5) | 67 (1.5) | 0.00 | 16 (1.6) | 9 (1.1) | 0.04 |
| [40%-50%) | 1,169 (16) | 778 (17) | 0.02 | 116 (12) | 95 (12) | 0.02 |
| [50%-60%) | 4,804 (68) | 3,023 (67) | 0.01 | 716 (72) | 556 (72) | 0.01 |
| [60%-82%] | 914 (13) | 534 (12) | 0.03 | 125 (13) | 99 (13) | 0.00 |
| Unknown | 113 | 9 | 0.12 | 19 | 1 | 0.13 |
| Chronic pulmonary disease | 449 (6.2) | 298 (6.6) | 0.02 | 87 (8.6) | 74 (9.5) | 0.03 |
| Peripheral arterial disease | 548 (7.6) | 369 (8.2) | 0.02 | 94 (9.4) | 80 (10) | 0.03 |
| Neurological dysfunction | 67 (1.0) | 48 (1.1) | 0.01 | 15 (1.5) | 11 (1.4) | 0.01 |
| Unknown | 218 | 76 | 0.09 | 37 | 14 | 0.11 |
| Critical preoperative state | 71 (1.0) | 63 (1.4) | 0.03 | 13 (1.2) | 13 (1.7) | 0.03 |
| Recent myocardial infarction | 2,287 (32) | 1,483 (33) | 0.03 | 308 (31) | 245 (32) | 0.02 |
| Unknown | 7 | 3 | 0.01 | 1 | 0 | 0.04 |
| Euroscore II | 1.40 ±<br>1.35 | 1.43 ± 1.52 | 0.02 | 2.38 ±<br>2.58 | 2.41 ± 2.50 | 0.01 |
| Unknown | 17 | 13 | 0.01 | 5 | 4 | 0.01 |
| Procedural urgency |  |  |  |  |  |  |
| Elective | 3,579 (50) | 2,225 (50) | 0.02 | 467 (47) | 378 (49) | 0.04 |
| Urgent | 3,409 (48) | 2,188 (49) | 0.01 | 491 (50) | 373 (48) | 0.03 |
| Emergent | 105 (1.5) | 70 (1.6) | 0.01 | 28 (2.9) | 21 (2.7) | 0.01 |
| Salvage | 9 (0.1) | 6 (0.1) | 0.00 | 5 (0.5) | 2 (0.2) | 0.05 |
| Unknown | 102 | 1 | 0.13 | 18 | 0 | 0.15 |
| Prior CVA | 176 (2.9) | 118 (3.1) | 0.01 | 33 (4.0) | 26 (3.9) | 0.01 |
| Unknown | 1211 | 616 | 0.09 | 181 | 106 | 0.12 |

Mean ± standard deviation, number (percentage). Abbreviations: CVA, cerebrovascular event; eGFR, estimated glomerular filtration rate; LVEF, left ventricular ejection fraction; RA, radial artery; RITA, right internal thoracic artery; SMD, standardized mean difference.

### Operative data

Off-pump surgery was more common in the RITA cohort than in the RA cohort for both women (38% vs 13%, p<0.001) and men (37% vs 11%, p<0.001). The use of an additional venous graft was more frequent in the RA cohorts (women 19% vs 8.3%, p=<0.001; men 24% vs 9.2%, p<0.001) (S2 Table).

### Clinical outcomes

Prior to inverse probability treatment weighting adjustment, the RITA cohort demonstrated higher mortality rates compared to the RA cohort in both men (p<0.001) and women (p<0.001) (S3 figure). After adjustment, survival did not differ significantly between treatment groups in both men (p=0.32) and women (p=0.72) (Fig 1).

**Figure 1.**
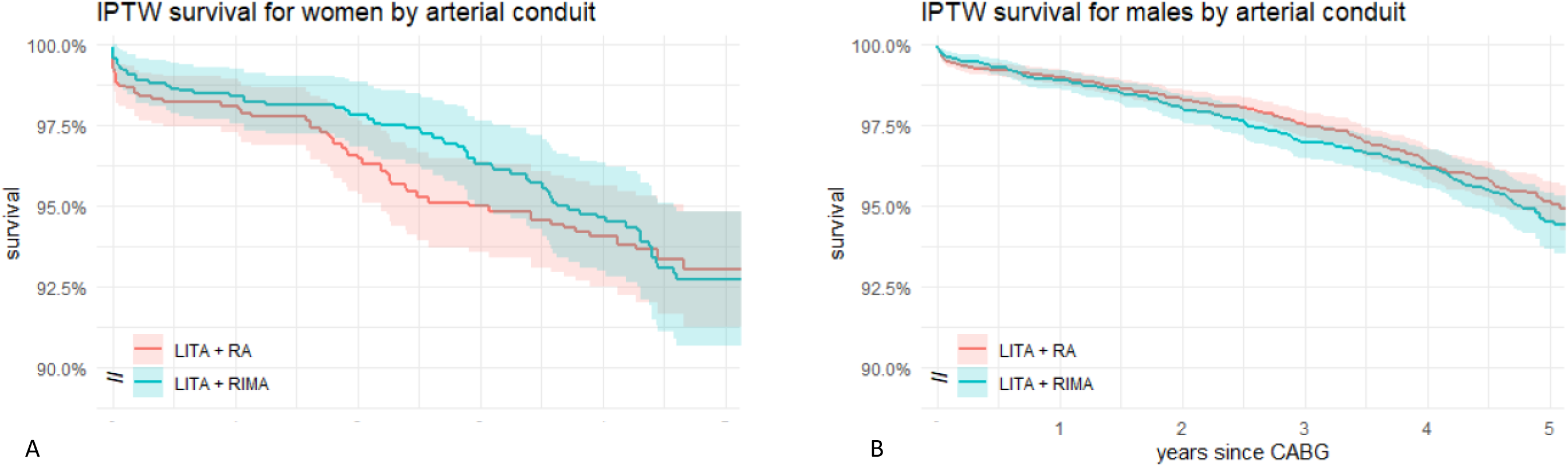
Kaplan Meier curve survival after inverse probability treatment weighting (IPTW) divided by arterial conduit for (A) women (p=0.72) and (B) men (p=0.32).

At 1-, 2-, and 5-year follow-up, the incidence of repeat revascularization remained similar between female cohorts (Figure 2; S4 Table). In the male population, the RA cohort exhibited a significantly higher risk of repeat revascularization at 2 and 5 years postoperatively (Figure 3; S4 Table).

**Figure 2.**
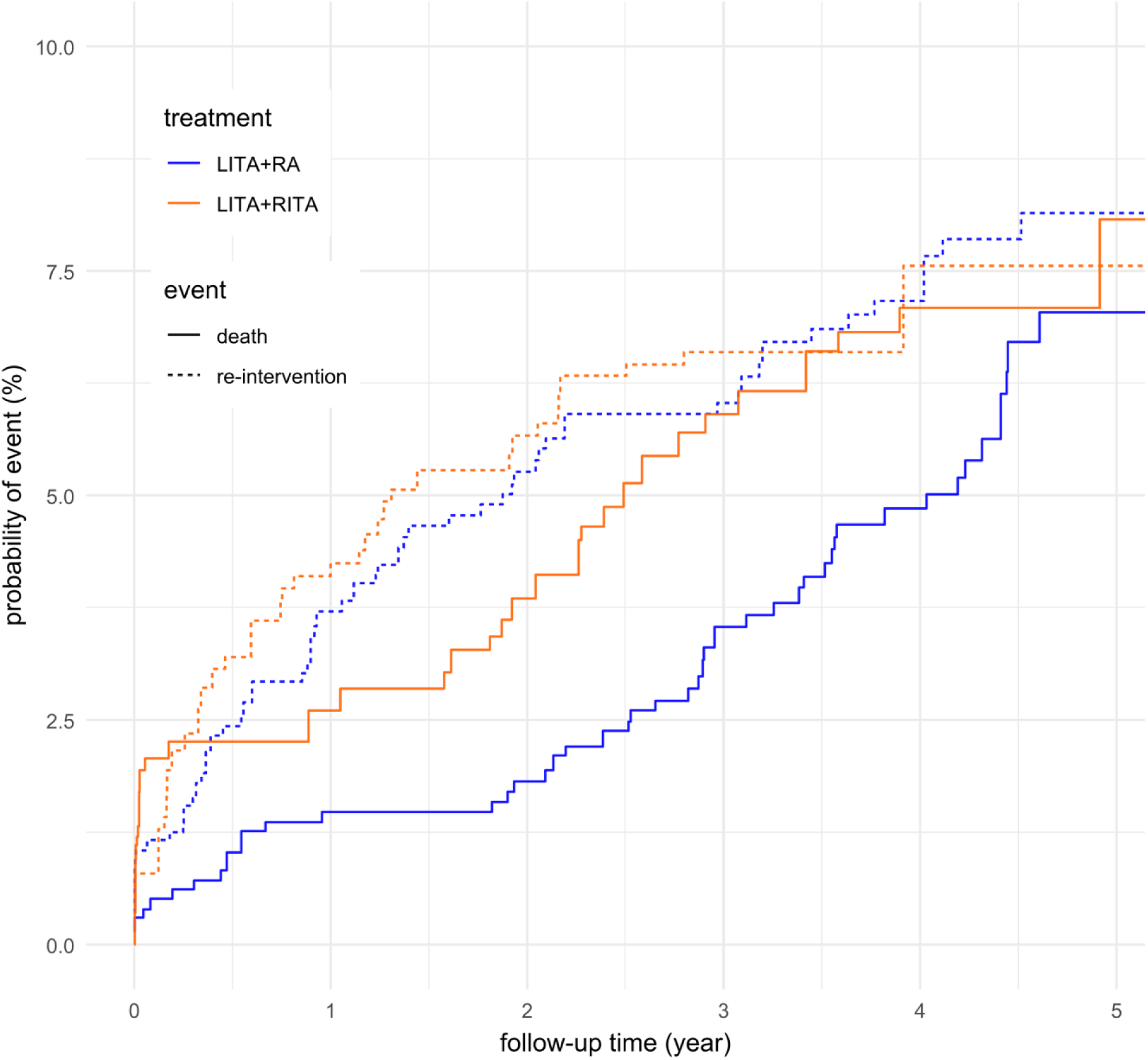
Competing risk analysis cumulative incidence plot of repeat revascularization and death during follow-up in women after inverse probability treatment weighting.

**Figure 3.**
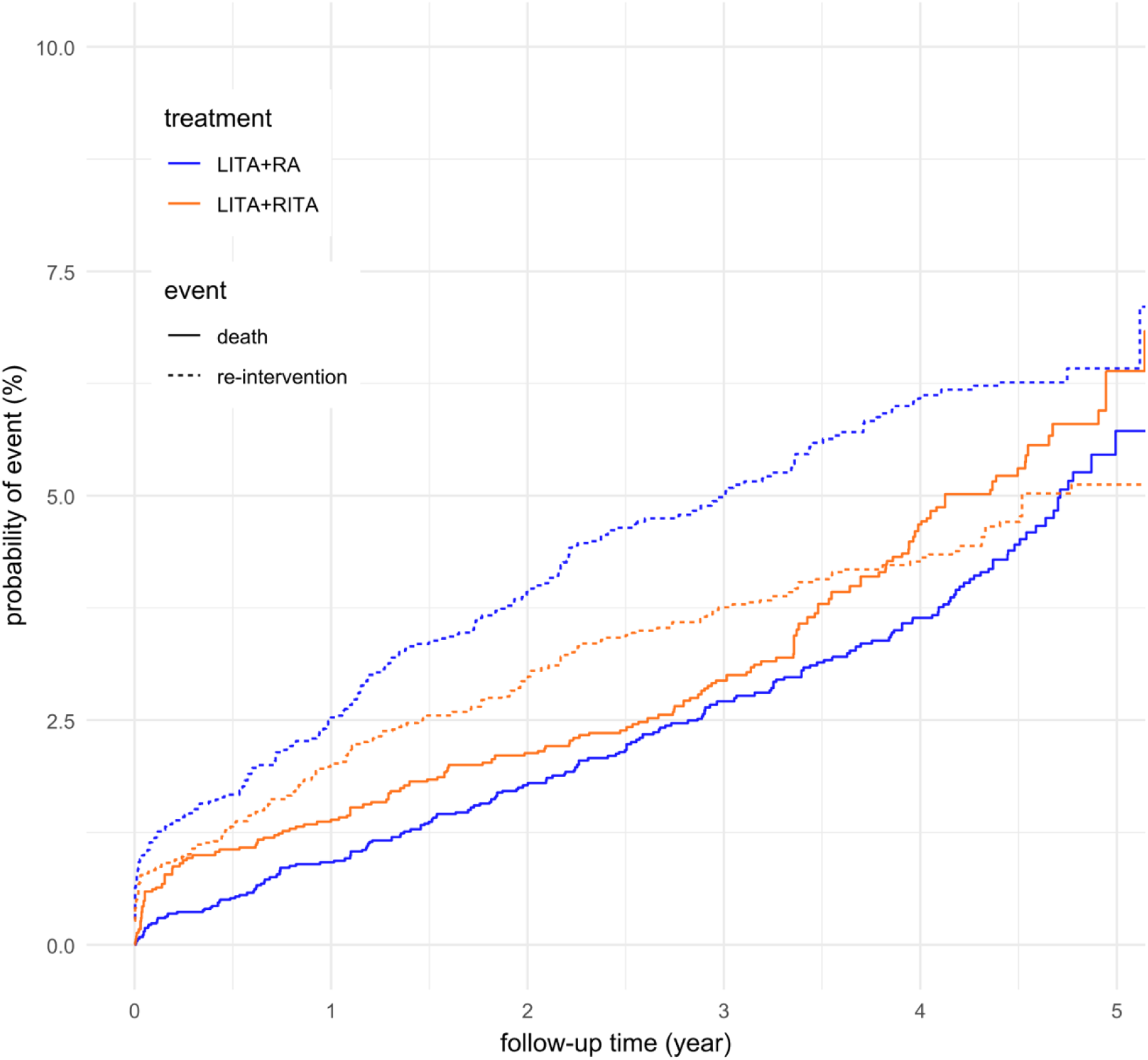
Competing risk analysis cumulative incidence plot of repeat revascularization and death during follow-up in men after inverse probability treatment weighting.

No significant differences in mortality or the need for repeat cardiac surgery during the postoperative hospital stay were observed after revascularization with the RA or the RITA. Among male patients, no difference was observed in the incidence of CVA between the RA and RITA cohorts. In the female population, CVA was more frequent in the RA cohort (0.9% vs. 0.2%, p=0.028). Conversely, use of the RITA was associated with a higher incidence of postoperative arrhythmias in men (20.5% vs. 16.4%, p<0.001) and women (20.1% vs. 15.2%, p=0.004) (table 3).

**Table 3.** Short-term postoperative complications. Number (percentage).

|  | **Men** |  |  | **Women** |  |  |
| --- | --- | --- | --- | --- | --- | --- |
|  | **RA** | **RITA** | **p-value** | **RA** | **RITA** | **p-value** |
|  | N = 7,204 | N = 4,490 |  | N = 1,009 | N = 774 |  |
| Death | 44 (0.6) | 23 (0.5) | 0.5 | 13 (1.3) | 6 (0.8) | 0.3 |
| Repeat cardiac surgery | 23 (0.3) | 23 (0.5) | 0.10 | 5 (0.5) | 2 (0.3) | 0.5 |
| CVA | 16 (0.2) | 13 (0.3) | 0.6 | 9 (0.9) | 1 (0.2) | **0.028** |
| Arrhythmias | 1,149<br>(16.4) | 912<br>(20.5) | **<0.001** | 149<br>(15.2) | 159<br>(20.1) | **0.004** |

Abbreviations: CVA, cerebrovascular event; RA, radial artery; RITA, right internal thoracic artery.

## DISCUSSION

Among both men and women undergoing isolated CABG, long-term mortality was comparable after MAG using the RA or RITA as the second arterial graft.

Women undergoing CABG generally experience higher mortality rates compared to men [6,11,12,15]. The underlying etiology of excess mortality observed in the female population following CABG has not been elucidated. Extensive research has demonstrated that women are at an increased risk of graft failure, which is independently associated with all-cause mortality [10,18]. Long-term graft patency and survival after MAG are superior compared to saphenous vein grafts in both sexes [4–6,19]. In the RAPCO-RITA trial, the 10-year graft patency rate and survival were significantly higher for the RA compared to the RITA. This benefit was observed in both sexes, although its magnitude appeared greater in women [9]. Similarly, a recent analysis of seven RCTs examining graft failure rates, showed 11.8% failure of the RA and 33.3% of the RITA in women, whilst corresponding rates in men were 14.1% and 21.6%. In our male population, the RA cohort had a higher rate of repeat revascularization [18]. This finding cannot solely be attributed to a more frequent use of additional vein grafts, as the female RA cohort did not exhibit a similar effect.

Histopathological studies comparing these two arterial conduits have shown increased susceptibility of the RA to intimal hyperplasia, atherosclerosis and medial calcification [13,20]. These pathological changes are especially pronounced in men, potentially contributing the increased rate of repeat revascularization observed during follow-up [20]. Although several studies have made an association between graft failure and mortality, we did not observe increased mortality in the male RA cohort despite the higher need for repeat revascularization.

Postoperative atrial fibrillation (POAF) is a common postoperative complication that significantly increases long-term mortality and morbidity [21–23]. Studies exploring the relationship of graft choice and POAF are limited. Local and systemic inflammation has been proposed as key contributor to the development of POAF [24]. The larger intrathoracic surgical wound associated with BITA use may lead to greater local inflammation and, consequently, an increased risk of POAF [25–27]. Caputo et al. reported more POAF in patients receiving a RITA compared to the RA [28]. Other known risk factors include on-pump CABG and revascularization by means of a single arterial graft with concomitant vein graft [29,30]. Despite higher rates of concomitant vein grafting and extracorporeal circulation use in the RA cohorts – known risk factors for POAF – we observed substantially more postoperative arrhythmias in the RITA cohorts. This finding may be consistent with the hypothesis of increased inflammatory burden in BITA use and subsequently increased risk of POAF. Nevertheless, these results should be interpreted with caution, as numerous other factors have been proposed in relation to POAF and as such a causal relationship with graft choice cannot be stated with certainty.

A significant difference in the incidence of perioperative CVAs was observed in the RA cohort of the female population. Advanced age and baseline comorbidities place women at increased risk of perioperative CVA. Additional risk factors include aortic manipulation during cardiopulmonary bypass and cross-clamping [31–34]. As on-pump surgery and vein graft use were more prevalent in the RA cohort, aortic manipulation and pre-existing comorbidities – including a history of CVA in 4% of women – may have contributed to the elevated incidence of CVA in this group [35].

### Limitations

As this was a retrospective study, the potential for selection bias and unmeasured confounding must be acknowledged. Findings are subject to variation based on surgeon preference and expertise. Incomplete revascularization is not reported in the registry and may influence mortality and repeat revascularization. Certain postoperative complications, including mediastinis, could not be assessed due to missing data from the earlier years in the dataset. Another limitation is the difference in conduit composition between groups. Patients in the RA group received more concomitant vein grafts, which are associated with lower long-term patency compared to arterial grafts. This may have contributed to the higher incidence of reintervention (CABG or PCI) in the male RA cohort. However, as data regarding the specific grafts involved in these reinterventions were not available, the underlying cause of graft failure could not be determined.

## CONCLUSION

The study presents real-world data on a substantial population of men and women undergoing MAG using the RA or RITA. When the surgeon decides the revascularization strategy, mortality is similar between treatment groups in both sexes, in contrast to findings from the only RCT performed to date on this topic. The female RA cohort experienced more perioperative CVAs, while the male RA cohort demonstrated a higher incidence of repeat revascularization. Large-scale prospective trials are warranted to evaluate the potential benefit of sex-specific tailoring of grafting and operative strategy to optimize long-term outcomes after MAG – particularly for women, who remain underrepresented in cardiovascular research.

## Availability of data and materials

The data that support the findings of this article are available from the Netherlands Heart Registration (<u>NHR</u>) upon reasonable request. Most of the data collected by the NHR is publicly available on www.hartenvaatcijfers.nl

## Data Availability

The data that support the findings of this article are available from the Netherlands Heart Registration (NHR) upon reasonable request. Most of the data collected by the NHR is publicly available on www.hartenvaatcijfers.nl

## Abbreviations

BITA: bilateral internal thoracic arteries
CABG: coronary artery bypass grafting
CVA: cerebrovascular accident
eGFR: estimated glomerular filtration rate
LITA: left internal thoracic artery
LVEF: left ventricular ejection fraction
MAG: multi-arterial grafting
NHR: Netherlands Heart Registration
POAF: postoperative atrial fibrillation
RA: radial artery
RCT: randomized controlled trial
RITA: right internal thoracic artery
SMD: standardized mean difference

## Supporting information

S1 File. Members of the Cardiothoracic Surgery Registration Committee of the Netherlands Heart Registration.

S2 Table. Operative variables. Number (percentage).

S3 Figure. Crude survival divided by sex and graft type before inverse probability treatment weighting.

S4 Table. Competing risk analysis of repeat revascularization and mortality at 1-, 2- and 5-year follow-up.

