## Supplementary material for "The optimal second arterial graft and sex differences in coronary bypass surgery: 10-year national registry results": S1 file, S2 table, S3 figure, S4 table

S2 Table. Operative variables. Number (percentage).

|  | Men | | | Women | | |
| --- | --- | --- | --- | --- | --- | --- |
|  | **RA**  N = 7,204 | **RITA**  N = 4,490 | **p-value** | **RA**  N = 1,009 | **RITA**  N = 774 | **p-value** |
| Off-pump surgery | 792 (11%) | 1640 (37%) | **<0.001** | 132 (13%) | 297 (38%) | **<0.001** |
| Venous graft | 1,699 (24%) | 414 (9.2%) | **<0.001** | 191 (19%) | 64 (8.3%) | **<0.001** |

Abbreviations: LITA, left internal thoracic artery; RA, radial artery; RITA, right internal thoracic artery.

S3 Figure.

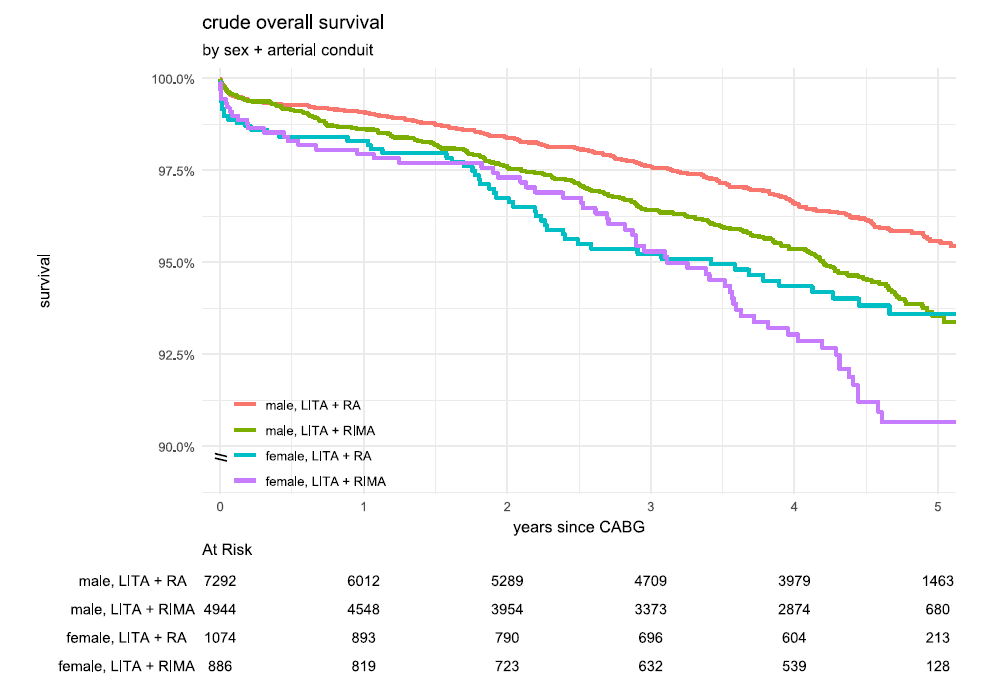

Crude survival divided by sex and graft type before inverse probability treatment weighting. Men, p<0.001; women, p<0.001. Abbreviations: LITA, left internal thoracic artery; RA, radial artery; RITA, right internal thoracic artery.

S4 table. Competing risk analysis of repeat revascularization and mortality at 1-, 2- and 5-year follow-up.

|  | Men | | | Women | | |
| --- | --- | --- | --- | --- | --- | --- |
| Time-point (years) | RD (%) | 95% CI | p-value | RD (%) | 95% CI | p-value |
| Repeat revascularization |  |  |  |  |  |  |
| 1 | -0.5% | -1.2% – 0.1% | 0.106 | +0.5% | -1.7% – 2.9% | 0.653 |
| 2 | -1.0% | -1.8% – -0.1% | **0.020** | +0.4% | -2.2% – 3.1% | 0.763 |
| 5 | -1.3% | -2.6% – 0.0% | **0.044** | -0.6% | 3.92% – 3.1% | 0.741 |
| Death |  |  |  |  |  |  |
| 1 | +0.5% | 0.0% – 1.0% | 0.072 | +1.1% | -0.4% – 3.0% | 0.919 |
| 2 | +0.4% | -0.3% – 1.0% | 0.283 | +2.0% | +0.2% – 4.2% | **0.043** |
| 5 | +0.7% | -1.0% – 2.5% | 0.445 | +1.0% | -2.5% – 5.0% | 0.585 |

Abbreviations: RD, rate difference; CI, confidence interval.
